# Using proper mean generation intervals in modelling of COVID-19

**DOI:** 10.1101/2021.03.25.21254307

**Authors:** Xiujuan Tang, Salihu S. Musa, Shi Zhao, Daihai He

## Abstract

In susceptible-exposed-infectious-recovered (SEIR) epidemic models, with the exponentially distributed duration of exposed/infectious statuses, the mean generation interval (GI, time lag between infections of a primary case and its secondary case) equals the mean latent period (LP) plus the mean infectious period (IP). It was widely reported that the GI for COVID-19 is as short as 5 days. However, many works in top journals used longer LP or IP with the sum (i.e., GI), e.g., > 7 days. This discrepancy will lead to overestimated basic reproductive number, and exaggerated expectation of infectious attack rate and control efficacy, since all these quantities are functions of basic reproductive number. We argue that it is important to use suitable epidemiological parameter values.

## Introduction

Emerging and re-emerging infectious diseases pathogens remain an enormous issue for public health and socio-economic growth because they can spread rapidly worldwide. Coronavirus disease 2019 (COVID-19), a respiratory disease, caused by the severe acute respiratory syndrome coronavirus 2 (SARS-CoV-2) (Li et al., 2020; WHO, 2021), has been a tremendous public health problem affecting every corner of the world (WHO, 2021). Since its appearance in late 2019, about 124 million people contracted and over 2.7 million died worldly as of 25 March 2021 (WHO, 2021). Until recently, many clinical features and underlying etiology of the SARS-CoV-2 remain unclear. Timely treatment and effective non-pharmaceutical interventions (NPIs) control measures against disease is important for timely mitigation (WHO, 2021).

Generation interval (GI), which can also be referred to as the generation time, is the time lag between infection incidents in an infector-infectee pair (Svensson, 2007). It is a proxy of serial interval (SI) of infectious disease, which is the time lag between onsets of the symptoms in an infector-infectee pair (Ali et al., 2020). The SI and GI are vital biological quantities (epidemic parameters) used for estimating the basic reproductive number (denoted by *R*_0_) which is defined as the number of secondary cases that one infected person will generate on average over the course of his/her infectious period in a population that is completely susceptible (Li et al., 2020; Musa et al., 2020a), as well as time-varying basic reproduction number, *R*_0_ (*t*), which determines the average number of secondary cases per infectious case in a population made up of both susceptible and non-susceptible hosts (Ali et al., 2020). Moreover, the importance of GI is also reflected in the renewal equation 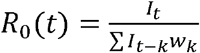, where *w*_*k*_ represents the GI distribution, *I*_*t*_ denotes daily infections and *R*_0_ (*t*) represents the daily instantaneous reproductive number, which reflects transmission dynamics at a time, *t* (Park et al., 2020). Recently, many works have been done to understand and/or estimate the GI, and its proxy, i.e., SI, associated with infectious diseases, including the SARS-CoV and the SARS-CoV-2 (Ali et al., 2020; He et al., 2020b; Lipsitch et al., 2003; Nishiura et al., 2020; Svensson, 2007; Wallinga and Lipsitch, 2007; Wang et al., 2020; Zhang et al., 2020; Zhao et al., 2020b).

Previous reports highlighted that when the SI is larger, the uncertainty and overestimation will be higher (Ali et al., 2020; Zhao et al., 2020a). The SI (which depends hugely on the incubation period of infectious disease) can be negative if the start of the symptoms in the infectee happens earlier than the start of the symptoms in the infector (person who transmit the disease) (Du et al., 2020; Ganyani et al., 2020a; Kong et al., 2020; Ren et al., 2020; Tindale et al., 2020; Zhao, 2020). The SI can also be negative when the incubation period has a wider range than the latent period, which may result in pre-asymptomatic transmission as reported in recent COVID-19 studies (Ali et al., 2020; He et al., 2020b). However, unlike SI, the GI is solely non-negative according to its definition (Wallinga and Lipsitch, 2007; Yan, 2008).

The incubation period is the time between infection and the onset of symptoms (Yan, 2008). Although the time of exposure for an individual who transmits the disease (infector) is usually indistinguishable, the time of exposure of an individual who gets the infection (infectee) can be determined by the contact tracing history of the ‘infector-infectee’ pair. This subsequently indicates that for ‘infector-infectee’ pairs, there is a single infector that relates to the infectee epidemiologically. Hence, the incubation periods of infectees may be identifiable. However, the latent period differs from the incubation period, it is defined as the time lag between the infection in exposure and onset (beginning) of infectiousness of a typical case (Yan, 2008). Since the beginning of infectiousness is indistinguishable, the latent period is unidentifiable. Thus, we noticed that in many diseases (mostly infectious), the mean latent period is less than or equal to the mean incubation period (such as COVID-19) (Ali et al., 2020), whereas some diseases have a long latent period, e.g., Ebola virus disease. Note that people infected with Ebola are not infectious until the symptoms started, the incubation period of Ebola varies between 2-21 days).

Moreover, the latent period is the time interval when an infected individual is unable to transmit the disease. While the time interval during which an infected individual can transmit the disease is called the infectious period. Both are random variables and are considered independent, thus, the LP and IP are not generally traceable. However, SI is identifiable and well-studied, and reported by epidemic models (Lipsitch et al., 2003; Nishiura et al., 2020). We observed that some studies in the literature did not use the LP and IP appropriately, as the sum of their mean equals to the mean generation interval in SEIR-based models, that is, mean GI = mean LP + mean IP. Using the same notation as in (Svensson, 2007), we have that, the expectation of the random generation interval is given by *E* (*T*) = *E* (*X*) + *E* (*Y*), where *T* is the random variable representing the generation interval of the infection, and *X* and *Y* represent the random latent time and random infectious time, respectively. Details on this relation in the SEIR-based model can be found in Svensson (Svensson, 2007).

Furthermore, it is of vital importance to forecast the size of the outbreak, including infection attack rate (AR), the need for ventilators and hospital beds, the expected severe cases and deaths, the herd immunity threshold, and the vaccine supply needed. All of these are associated with the time-varying basic reproductive number, *R*_0_ (*t*). Given the important role of *R*_0_ (*t*), it is imperative to obtain their estimation more accurately. Therefore, it is crucial to use the proper value for the mean LP and mean IP in SEIR compartmental models. In this work, we showed that longer IP or LP leads to an overestimation of reproductive number, using COVID-19 confirmed death cases data for Belgium, Israel, and the United Arab Emirates (UAE). We noticed that Belgium was hit badly by two waves. Israel and UAE have started large-scale vaccination programs. We choose these countries (as an example) to demonstrate the impact of the GI on *R*_0_ (*t*) to provide more qualitative insights on the use of the GI for controlling the disease outbreaks.

## Methods

We use the COVID-19 reported deaths data retrieved from the official website of the World Health Organization (WHO) public surveillance reports for Belgium, Israel, and UAE available from https://covid19.who.int/ (WHO, 2021). The time-series distribution of weekly confirmations of COVID-19 cases and deaths in Belgium, Israel, and UAE were depicted in **Figure 1** which shows the patterns of the COVID-19 epidemics in these three countries. The cases and deaths for COVID-19 are represented by black and red dotted curves respectively. We observed that Israel and UAE show similar epidemic curve patterns. While Belgium was hit harder with the two waves of COVID-19 outbreaks. The population data for the three countries were obtained from the Worldmeter, available from https://www.worldometers.info/population/ (WM, 2021).

**Figure 1:**
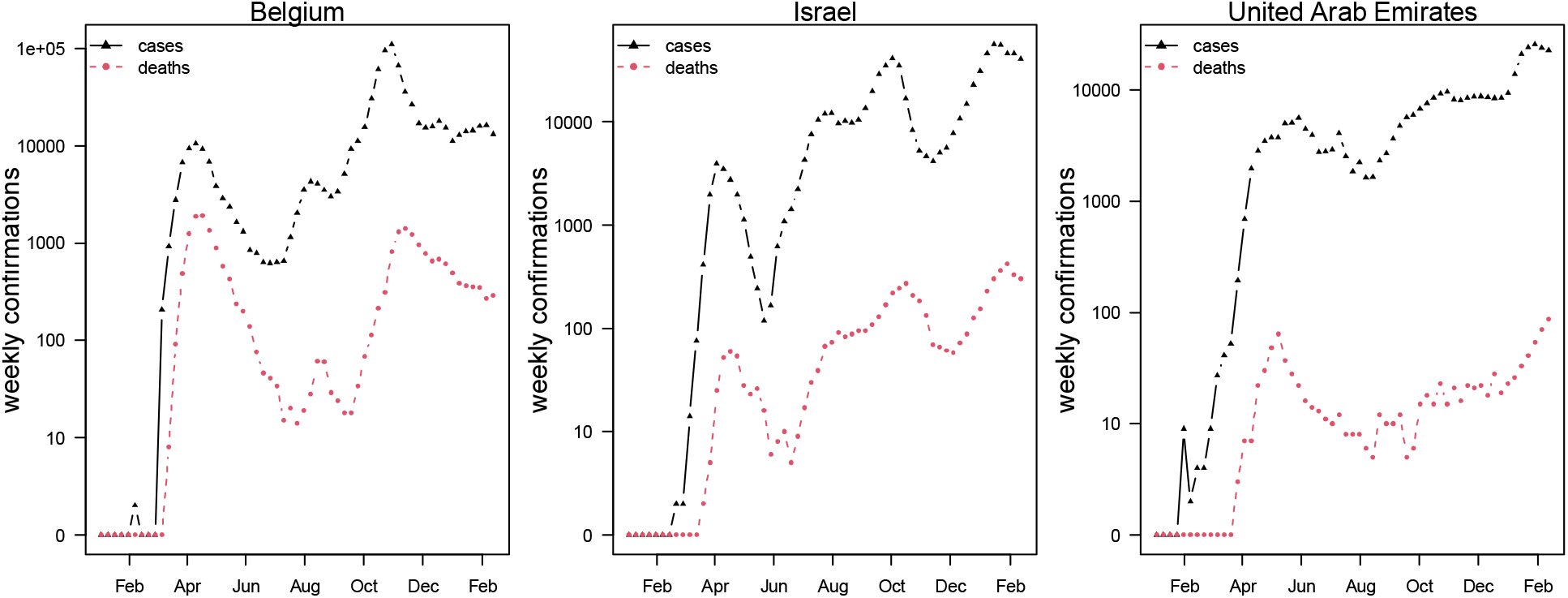
Weekly confirmed cases (in black triangles) and deaths (in red tringles) of COVID-19 in Belgium, Israel, and the United Arab Emirates.

Thus, we formulate the following simple epidemic model.

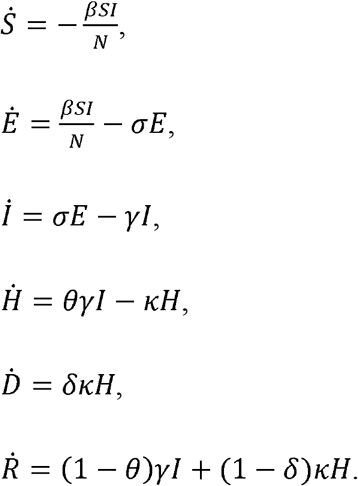

Here, *S, E, I, R, H*, and *D* represent susceptible, exposed, infection, recovered, hospitalized, and death classes. The parameters *β, σ, γ,k* are transmission rate, progression rate from *E* to *I*, recovery rate (for fitting simplicity, we assumed the recovery rate and hospitalization rate to be the same), the proportion of individuals moving from *H* to *D*, respectively. *θ* represents proportion of hospitalization (or recorded cases or severe cases) among infection and *δ* represents the proportion of death among hospitalization (or recorded cases or severe cases). Here, we assumed that hospitalization can be interpreted as symptomatic cases. Since we do not explicitly incorporate the hospitalization cases in fitting, the exact definition is not crucial. The infection fatality rate equals *δθ*. We consider three scenarios on the choice of *δ*, including *δ* = *θ, δ* = 0.1, and *δ* = 0.05. We fit the daily integrated D to the reported deaths in each country. We assume a negative nominal measurement noise in reporting with an over-dispersion parameter *τ*. We assume a time-varying *β*, which is an exponential cubic spline function with the number of nodes as 7, which was evenly distributed over the study period from 1 March 2020 to 18 Feb 2021. The time-varying basic reproductive number is given by *R*_0_ (*t*) *≈β*(*t*)/ *γ*. The detailed model-fitting method can be found in many previous studies (He et al., 2020a; Musa et al., 2020b; Zhao et al., 2018).

In the classic susceptible-exposed-recovered-based models, the mean GI of an infectious disease equals the sum of the mean latent period (LP) and the mean infectious period (IP) (Svensson, 2007). The duration of individuals in an exposed/infectious class follows exponential distributions. Due to the discrete-time in the simulation of the model, the realized (or simulated) mean LP and mean IP according to He et al. (He et al., 2010) are, 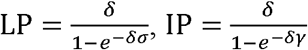, where *δ* designate the time discretization step. Thus *σ*^-1^ and *γ*^-1^ are theoretical mean LP and IP. The simulated periods were slightly larger than theoretical values due to the time discretization. The discrepancy diminishes when the time step size approaches zero. Hence, the sum of the mean LP and mean IP is estimated at 6.07 days with a 1-day time step size and theoretical 2 days LP and 3 days IP. The mean GI equals 5 days when the time step size approaches zero. Besides *σ*^-1^ at 2 days and *γ*^-1^ at 3 days, we set *k*^-1^=14 days and *θ*^*2*^ in the range of 0.5% to 1% (Mellan et al., 2020). All these petameter values are biologically reasonable.

Therefore, using iterated filtering methods, we fitted a SEIRD model with an additional death class to reported COVID-19 deaths in the three countries (i.e., Belgium, Israel, and UAE) to reveal the effects of the mean LP and the mean IP on the estimation of reproduction number. We fitted the model to COVID-19 deaths data since COVID-19 mortality data seems less affected by testing policy compared to other diseases.

## Results and discussion

Based on a recent study on GI and SI, we observed that most studies showed that the SI (and/or GI) of COVID-19 varies between 5-6 days (Griffin et al., 2020). In particular, Ferretti et al. (Ferretti et al., 2020) reported the mean GI as 5.0 days, Ganyani et al. (Ganyani et al., 2020b) used the data for Singapore and Tianjin, China, and found that the mean GI is estimated at 5.20 (3.78 – 6.78) days and 3.95 (3.01 – 4.91) days, respectively. In forty research papers reviewed by Griffin et al. (Griffin et al., 2020) on the GI and SI, only three studies provided an estimate for the mean GI, which varies roughly between 3.95 to 5.20 days. And one paper provided an estimate for the median of the GI as 5.0 days (Backer et al., 2020; Ferguson et al., 2020; Griffin et al., 2020; Li et al., 2020). Furthermore, Zhang et al. (Zhang et al., 2020) reported that the incubation period of COVID-19 was estimated at 5.2 (95%CI: 1.8-12.4), and the mean IP at 4.4 (95%CI: 0.0-14.0) from Dec 24 to Jan 27, 2020, and 2.6 (95%CI: 0.0-9.0) from Jan 28 to Feb 17, 2020.

However, several studies reported the period of disease progression before the infectiousness stage as the LP in an SEIR epidemic model. For example, Yin et al. (Yin et al., 2021) conducted a modelling study to assess the effectiveness of NPIs measures (including contact tracing, facemask wearing, and rapid testing) to curtail the spread of the COVID-19 in China. They reported that asymptomatic patients lasted 4.6 days in LP and 9.5 days in IP until removal. See **Table 1** for more details. Many studies did not follow the rule that mean GI= mean LP + mean IP < 6 days. Therefore, we emphasized that appropriate use of the generation interval in an epidemiological study is essential to effectively control the COVID-19 outbreaks, because it provides a more accurate estimate on reproduction number for the epidemics, and is crucial for pandemic mitigation planning and forecasting.

**Table 1:** Mean latent period and mean infectious period of COVID-19.

| Mean LP (days)/ Mean IP (days) | Equivalent Mean GI (days) | References |
| --- | --- | --- |
| None | 5.0 | (Ferretti et al., 2020) |
| None | 5.20 (3.78-6.78) for Singapore<br>& 3.95 (3.01-4.91) for Tianjin, China | (Ganyani et al., 2020b) |
| 5.2 (95%CI: 1.8-12.4) (incubation period)/ 4.4 (95%CI: 0.0-14.0) from Dec 24 to Jan 27, 2020, and 2.6 (95%CI: 0.0-9.0) from Jan 28 to Feb 17, 2020 | >5.2 | (Zhang et al., 2020) |
| 4.6/9.5 | 14.1 | (Yin et al., 2021) |
| 4.6/5 | 9.6 | (Kissler et al., 2020) |
| 4.3/(5+2.1+2.9=10) | 10 | (Emery et al., 2020) |
| 5.1 (incubation period) 12 (95% CI: 2-14) | >12 | (Lauer et al., 2020) |

For demonstration purposes, we compared epidemiologic dynamics of COVID-19 for some randomly selected countries (Belgium, Israel, and UAE) while varying LP and IP from (2 & 3 days to 3 & 6 days). We employed the model to the COVID-19 mortality data and obtained the time series fitting results using the COVID-19 data for Belgium, Israel, and UAE to quantify the effects of longer GI on the estimation of (time-varying) reproduction numbers. In **Figure 2** we show the fitting results of the daily confirmed COVID-19 deaths (red circled) with proper LP and IP at 2 & 3 days. in (a) Belgium, (b) Israel, and (c) UAE, respectively. The median of the simulation is represented by the black curve, and the time-varying basic reproduction number is denoted by the blue dashed curve. The 95% range of the simulation is shown by the shaded gray region. In **Figure 3**, we compare six scenarios, corresponding *δ* = *θ, δ* = 0.1, and *δ* = 0.05, with proper GI (LP and IP at 2 & 3 days) versus long GI (LP=3 days and IP=6 days) for each country. From **Figure 3**, we discovered that using longer mean LP and mean IP would significantly increase an estimate of reproduction number. Thus, the magnitude of reduction in the initial reproduction number would be much higher in the latter cases (3&6 days for LP&IP) than in the proper former cases (2&3 days for LP&IP). Besides, a higher initial reproduction number would imply a much higher expected infection attack rate and herd immunity threshold.

**Figure 2:**
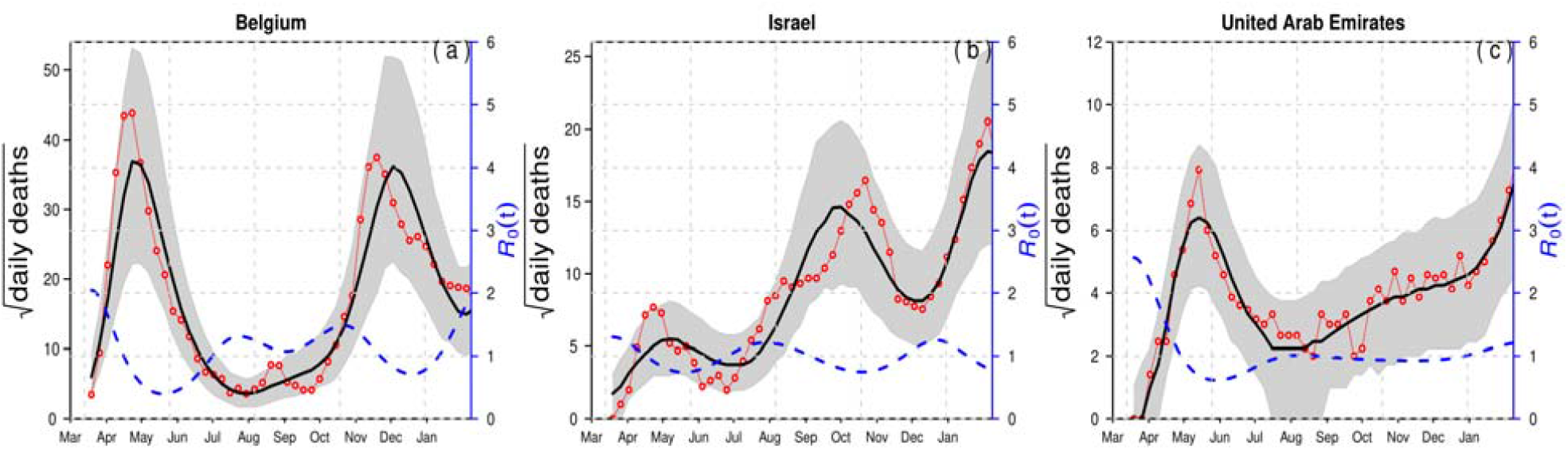
Time series fitting results of daily confirmed COVID-19 deaths (in red circled) in (a) Belgium, (b) Israel and (c) the United Arab Emirates represented, respectively. The medium of the simulation is represented by the black curve, and the time varying reproduction number (*R*_0_ (*t*)) is denoted in blue dashed curve. The 95% confidence interval of the simulation is shown by the shaded (gray) region. The mean LP = 2 days and the mean IP = 3 days. Thus, the mean GI = 5 days.

**Figure 3:**
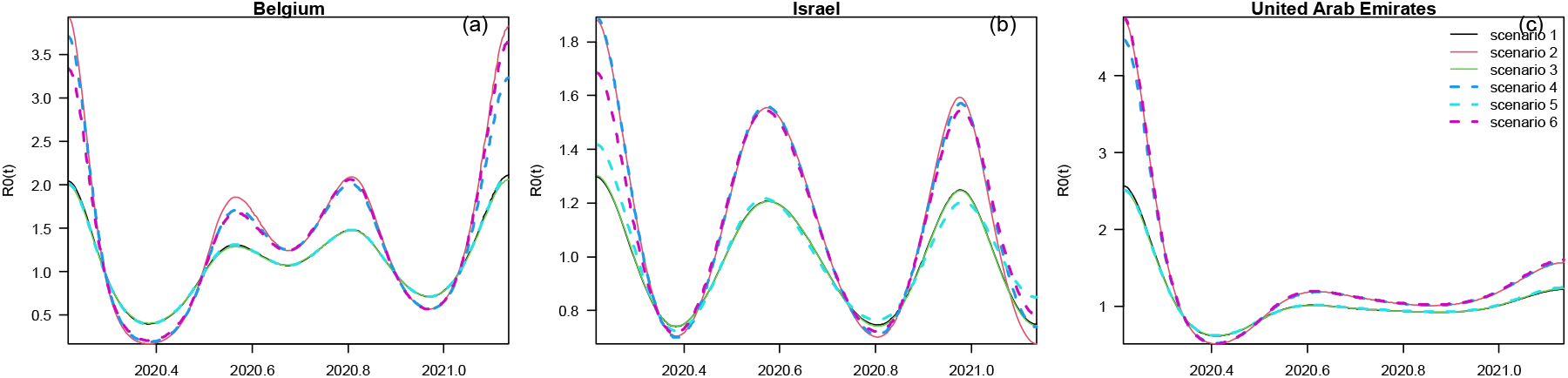
Comparison of reconstructed R0(t) under six different scenarios for three countries (a) Belgium, (b) Israel and (c) the United Arab Emirates, respectively. The mean LP = 2 days and the mean IP = 3 days in scenarios 1,2,3. We assumed *δ* = *θ* in scenario 1&2, while we fix *δ* = 0.1 in scenario 3&4, and *δ* = 0.05 in scenario 5&6, respectively. The mean LP = 3 days and the mean IP = 6 days in scenarios 2,4,6, thus GI=9 days. In all countries, the reconstructed R0(t) with longer GI is evidently higher that with a shorter GI. If the initial drop of R0(t) is associated with control measure, the assessment with a longer GI will lead to overestimated control effectiveness.

Furthermore, a summary results of the COVID-19 infection attack rates (AR) for Belgium, Israel, and UAE is presented in **Table 2** with reasonable LP and IP values. The choice of LP and IP had an important influence on the estimate of AR, likely due to the choice of the flexible transmission rate in our model, and the assumption of the infection fatality rate which varies between 0.5% to 1%. **Appendix Table 1** presents the results of the estimated parameter values using the log-likelihood estimation approach for Belgium, Israel, and UAE. We observed that Belgium has the lowest log likelihood values, indicating that Belgium was hit harder than the other two countries. also, a summary results of the estimated values for the time-varying transmission rate with a fixed number of nodes (denoted by *n*_*m*_) was given in **Appendix Table 2**. The initial values for the state variables used for the model given in **Appendix Table 3**. Therefore, based on the results obtained and the comparison of the epidemic dynamics of COV-ID-19 for Belgium, Israel, and UAE with varying LP and IP, we argue that appropriate LP and IP should be used in epidemiological modelling study to effectively mitigate the spread of disease and to provide suitable suggestions of control strategies for public health implementation and policymaking.

**Table 2:** A summary results of the estimated infection attack rates (AR) in Belgium, Israel, and the UAE by February 18, 2021.

| Country | Population | Death | AR |
| --- | --- | --- | --- |
| Belgium | 11589623 | 21041 | 0.182 |
| Israel | 8655535 | 4634 | 0.059 |
| UAE | 9890402 | 819 | 0.009 |

## Conclusions

Mean LP, IP, and GI are essential quantities in epidemiological modelling studies that are used for the estimation of reproductive number of an infectious disease. For COVID-19, current knowledge showed that the mean GI (mean LP + mean IP) varies between 5-6 days, which implies that the mean LP/IP in SEIR models should be around 2-3 days, respectively. We emphasized that this estimate should be used to provide a reasonable estimation of the reproductive number (*R*_0_), which helps in policymaking to curtail the spread of an infectious disease. We showed that the estimated R_0_(t) for Belgium, Israel, and UAE are elevated substantially when longer LP and IP are used. Since now vaccination programs are ongoing in these countries, all modeling fitting is timely to lay the groundwork for the efficacy evaluation of the vaccination programs in these countries.

## Data Availability

All data used are publicly available.

## Declarations

### Ethical approval and consent to participate

Not applicable.

### Consent for publication

Not applicable.

### Availability of data and materials

All data used are publicly available.

## Author contributions

All authors contributed equally for this manuscript.

## Acknowledgements

Not applicable.

**Uncategorized References**

## Appendices

**Title:** Using proper mean generation intervals in modelling of COVID-19.

**File name:** Supplementary material 1 - Appendix.docx

**Title of data**: Appendix

**Description of data**: Contains the following.

**Appendix Table 1.** Summary table of estimation results of the parameters (using log likelihood).

| Country | loglik | loglik.sd | $\sigma$ | $\theta$ | $\gamma$ | $\kappa$ | Pop size |
| --- | --- | --- | --- | --- | --- | --- | --- |
| Belgium | -284.8281 | 0.0229 | 182.5 | 0.1 | 121.6667 | 26.0714 | 11589623 |
| Israel | -229.7767 | 0.034 | 182.5 | 0.0954 | 121.6667 | 26.0714 | 8655535 |
| UAE | -139.7897 | 0.0829 | 182.5 | 0.0933 | 121.6667 | 26.0714 | 9890402 |
| Belgium | -285.0237 | 0.026 | 121.6667 | 0.0935 | 60.8333 | 26.0714 | 11589623 |
| Israel | -230.729 | 0.039 | 121.6667 | 0.1 | 60.8333 | 26.0714 | 8655535 |
| UAE | -138.8795 | 0.0593 | 121.6667 | 0.0826 | 60.8333 | 26.0714 | 9890402 |

**Appendix Table 2.** Summary table of estimation results of the time varying transmission rate with number of nodes (*n*_*m*_ = 7) fixed.

| log.beta | log.beta1 | log.beta2 | log.beta3 | log.beta4 | log.beta5 | log.beta6 |
| --- | --- | --- | --- | --- | --- | --- |
| 5.5159 | 3.9121 | 4.9633 | 4.8712 | 5.1678 | 4.4799 | 5.5484 |
| 5.0615 | 4.5097 | 4.9234 | 4.8167 | 4.5318 | 5.0215 | 4.5114 |
| 5.7429 | 4.4287 | 4.6888 | 4.7855 | 4.7286 | 4.7858 | 4.9997 |
| 5.4773 | 2.3899 | 4.5058 | 4.3241 | 4.797 | 3.5729 | 5.4488 |
| 4.7398 | 3.7754 | 4.4478 | 4.2501 | 3.7927 | 4.5705 | 3.7169 |
| 5.6623 | 3.6152 | 4.0472 | 4.2476 | 4.1243 | 4.2418 | 4.5581 |

**Appendix Table 3:** Summary table for initial values of state variable of the model.

| S.0 | E.0 | I.0 | T.0 | D.0 | R.0 |
| --- | --- | --- | --- | --- | --- |
| 11010142 | 4547 | 4547 | 455 | 45 | 569887 |
| 8222758 | 392 | 392 | 39 | 4 | 431950 |
| 9395882 | 11 | 11 | 1 | 0 | 494496 |
| 11010142 | 7231 | 7231 | 723 | 72 | 564225 |
| 8222758 | 471 | 471 | 47 | 5 | 431783 |
| 9395882 | 22 | 22 | 2 | 0 | 494474 |

